# Policy perspectives on the sustainable introduction of pneumococcal conjugate vaccines in middle-income countries

**DOI:** 10.1101/2024.06.07.24308483

**Authors:** Rose Weeks, Baldeep K. Dhaliwal, Jasmine Huber, Sarah Nabia, Ala’a F. Al-Shaikh, Kulkanya Chokephaibulkit, LaKKumar Fernando, Ehssan Baghagho, Anita Shet

## Abstract

Despite the well-documented life-saving potential of pneumococcal conjugate vaccines (PCV) and global efforts to widen vaccine availability, access to PCV in middle-income countries (MICs) has remained suboptimal due, in part, to vaccine pricing and limited external funding opportunities. To understand gaps and opportunities for improving vaccine equality, this qualitative study engaged government policymakers and program leaders from MICs that do not currently have PCV in their national immunization programs to explore their perspectives on decision-making contexts and constraints related to PCV introduction. In-depth interviews with 17 participants from Egypt, Jordan, Sri Lanka, and Thailand in 2023 documented that, despite high routine immunization coverage and commitment toward building equitable health systems, implementing new vaccines such as PCV has remained challenging. Among the six thematic areas that emerged, two were strong enablers to vaccine implementation: 1) the existence of strong primary healthcare systems; and 2) established policy processes for vaccine decision-making. Three themes that emerged have historically hindered PCV introduction, including; 1) limited information on disease burden and available vaccine products; 2) competing country health priorities; and 3) financing challenges. The interplay of these thematic areas has documented a paradox unique to MICs, further contributing to inequalities in vaccine access. While a subset of MICs recently became eligible for support from Gavi, the Vaccine Alliance for introducing new vaccines, the marketplace has historically lacked tiered vaccine pricing that MICs could sustain for the long term. This is despite the great need with existing inequities and a substantial proportion of the world’s low-income and displaced populations. Finally, participants pointed to opportunities to address barriers through support from global and regional actors providing technical capacity-strengthening, advocacy, and strategic financial support. These findings are informative for strengthening equality in access to vaccines and developing sustainable strategies to introduce and sustain life-saving childhood vaccines, including PCV.

## Background

Considering the unmatched impact and cost-efficiency of immunization as a public health intervention, sustained investment in vaccination programs is imperative to achieve global objectives of reducing preventable diseases and mortality. However, recent trends indicate that certain country income groups, including middle-income countries (MICs), often face challenges in incorporating new vaccines into their immunization programs due in large part to financial constraints [1]. While 81% of low-income countries had introduced PCV in 2022, fewer than 70% of upper-middle-income countries (UMICs) had introduced PCV[2]. MICs predominately finance vaccines using their national budgets, as they are less able to rely on non-governmental organizations and foreign aid to assist in the facilitation of introducing new vaccines. Vaccine pricing available to MICs has varied substantially, with the bilaterally negotiated price of PCV for some MICs reported up to 12 times higher than the price available to other MICs[3]. The Pan American Health Organization (PAHO) has traditionally supported all member states (i.e., all countries in the Americas, and select participating and observing states in Europe) with PAHO’s Revolving Fund. The Revolving Fund allows member states to access vaccines at a heavily reduced rate through its pooled procurement mechanism[4]. Unfortunately, PAHO pricing has not been accessible to many MICs outside of the Americas.

The concerns that MICs pose to the global vaccine agenda are threefold; first, MICs dominate in numbers as the number of LICs has drastically declined due to global economic growth; second, MICs are home to three-quarters of the world’s population and 62% of the world’s poor; and third, MICs have slower uptake of new and priority vaccines, in part due to historical differences in pricing available to MICs for new vaccine introduction [5]. For the 2024 fiscal year, utilizing 2022 income data, the World Bank defines lower middle-income countries (LMICs) as economies with a gross national income (GNI) per capita of between US$ 1,136–4,465 and UMICs as those with a GNI per capita of between US$4,466–13,845[6]. These countries account for 67% of vaccine-preventable deaths[7], [8].

As such, without significant efforts to accelerate the pace of new vaccine introduction in MICs, the target of averting 50 million deaths globally by immunization during the Immunization Agenda 2030 (IA2030) decade (2021–2030) will not be met. With growing knowledge of these dynamic challenges, donors have expressed interest and made commitments to support vaccine introduction in MICs. One such donor is Gavi, the Vaccine Alliance (Gavi), which recently instituted a strategy called the ‘MICs Approach’ to address threats to the equity and sustainability of routine immunization programs in lower middle-income countries and IDA-eligible economies[9]. With a funding allocation of $300 million USD to support vaccine introduction in MICs for the 2021–2025 period, the MICs Approach aims to prevent backsliding in vaccine coverage and drive the sustainable introduction of PCV as well as rotavirus and human papillomavirus (HPV) vaccines, providing start-up vaccine financing for eligible countries equal to half of the first target cohort[10]. Delays in the introduction of these three vaccines have hindered countries’ ability to drastically reduce deaths caused by these three preventable diseases. While this new commitment has created opportunities for 46 LMICs to become eligible for time-limited, catalytic Gavi support to introduce new vaccines for children in their countries, UMICs continue to have little or no access to external funding for the implementation of new vaccines[11].

Although the World Health Organization (WHO) has recommended since 2007 that all countries should include PCV in their national immunization schedules to prevent life-threatening pneumonia, meningitis, and other pneumococcal diseases, as of 2022, 39 WHO member states have yet to introduce PCV into either their national or subnational immunization programs[12], [13].

Recognizing the limited evidence from the policy perspective on immunizations in MICs, particularly considering COVID-19-related pressures on immunization systems and ongoing investments in public health interventions, this study aimed to understand the determinants of PCV introduction in Egypt, Jordan, Sri Lanka, and Thailand. All but Thailand, a UMIC, are countries newly eligible for support from Gavi’s MICs Approach. Through this study, we aim to provide insight on the enablers and challenges for PCV introduction in MICs, which can in turn inform strategies for vaccine introduction and sustainability.

## Methods

### Study Design and Description

This qualitative study leveraged purposive sampling to identify key stakeholders in MICs who are critically involved in PCV decision-making. Once identified, trained researchers conducted semi-structured interviews to elicit participant views on unique factors in MICs that are likely to influence successful PCV introduction, rollout, uptake, and acceptance on a national scale. Tool development, data collection, and analyses were completed by the end of August 2023.

### Ethics

Study details and expectations were relayed to participants before interviews began. Prior to initiating interviews, participants provided oral informed consent after discussing study details. The Institutional Review Board at the Johns Hopkins Bloomberg School of Public Health determined this study to be exempt from human subjects’ research oversight (IRB #21009).

### Participants

The target population for this study were implementation leaders, including immunization program managers at Ministries of Health (MOH), as well as in-country technical experts from United Nations (UN) agencies, research institutions, and National Technical Advisory Groups on Immunization (NITAG). These participants are considered ‘elite’ interviewees, meaning individuals with substantial expertise in the topic of interest. Their perspectives are highly relevant, as they are typically in a position of power to facilitate large-scale change[14]. We used purposive sampling to initially identify participants with substantial expertise, and we used snowball (chain) sampling to further identify participants.

### Semi-structured interviews

The semi-structured interview guide used to conduct interviews included open-ended questions on topics including, but not limited to, perceptions of disease burden and susceptibility, health priorities, decision-making processes, and implementation processes. These domains were designed based on a framework used in previous research studies, which was initially designed to understand vaccine policy decision-making and to develop strategies to support evidence-informed vaccine decision-making[15]. This framework was further tailored to align with research objectives and research questions. Interviews were conducted virtually over video or audio-conferencing platforms and lasted between 20 and 60 minutes. From a theoretical perspective, this research aimed to ‘study up’ to limit unequal power dynamics between the interviewer and interviewee as much as possible, as well as better understand the conditions that facilitate vaccine decision-making [16]. This approach has been used to understand vaccine introduction dynamics in countries afflicted by humanitarian crises[17].

### Data management and analysis

Interview recordings were transcribed using Temi (New York, NY) or Otter.ai (Mountain View, CA), and the interviewers manually checked transcripts against the recordings of the interviews to ensure accuracy. Two individuals analyzed the transcripts separately using the framework method, a qualitative analysis approach designed to rapidly facilitate the systematic and rigorous analysis of qualitative data[18]. Analysts initially started with the previously designed framework to guide matrix development[15]. Analysts further allowed the data to deductively guide the analysis process and support the expansion of this matrix. The matrix topics were finalized after several rounds of iterative reviews. The team then reviewed interview transcripts and extracted data into a matrix for each participant. These matrices also included a summary of participant views for each topic. By extracting data from each chart summary, the research team created an overall summary matrix, highlighting salient findings across all respondents. The summary matrix allowed the team to compare emerging themes as well as to facilitate comparisons between respondents and respondent groups (i.e., between countries). Lastly, data were condensed to highlight the core findings for each topic.

## Results

### Participants

There were 53 recruited individuals in the four countries^1^, 17 of whom were interviewed. Of these, three were based in Egypt, four in Jordan, five in Sri Lanka, and five in Thailand. Their characteristics are shown in Table 1.

**Table 1:** Participant Details.

| Country | Interviews | Sex (Male, M; Female, F) | Participants |
| --- | --- | --- | --- |
| Egypt | 3 | 3 F | Researcher (2), UN Agency (1) |
| Jordan | 4 | 4 M | MOH (2), Researcher (1), UN Agency (1) |
| Sri Lanka | 5 | 5 M | Researcher (2), Pediatric (3) |
| Thailand | 5 | 2F, 3M | MOH (1), Pediatric (1), Researcher (3) |

*Participant types: Ministry of Health representative (MOH), independent pediatric leader (Pediatric), research scientist (Researcher), and representative of UN agency (UN Agency)*

### Findings

Participants discussed six themes of importance for the introduction and implementation of PCV. Among these, two of the themes serve as strong enablers to vaccine implementation: strong primary healthcare systems; and established policy processes for vaccine decision-making. However, three of the themes were identified as obstacles to PCV introduction: limited information on disease burden and available vaccine products; competing country health priorities; and financing challenges. The last theme pointed to an opportunity: a role for increased support from global and regional actors via the provision of technical assistance, advocacy, or short-term financial support. These findings are detailed below and summarized in Table 2.

**Table 2:**
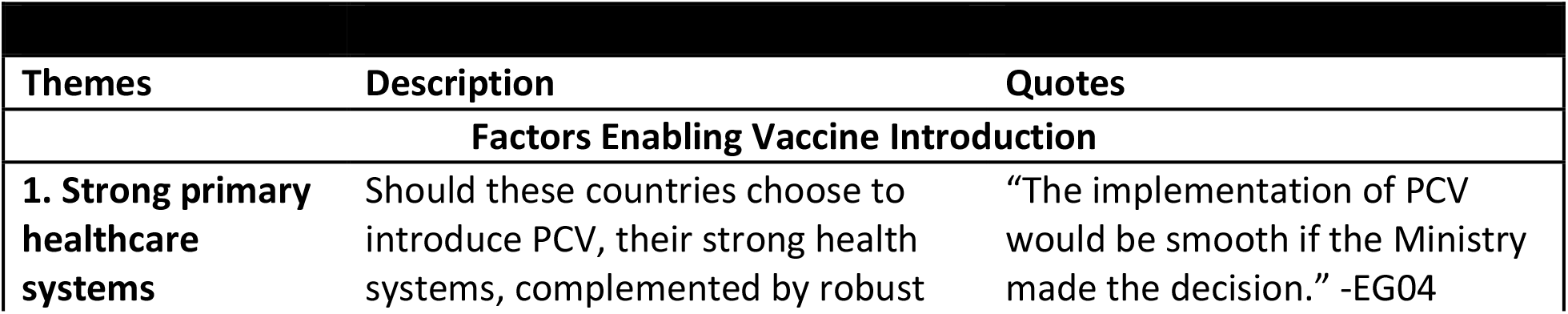
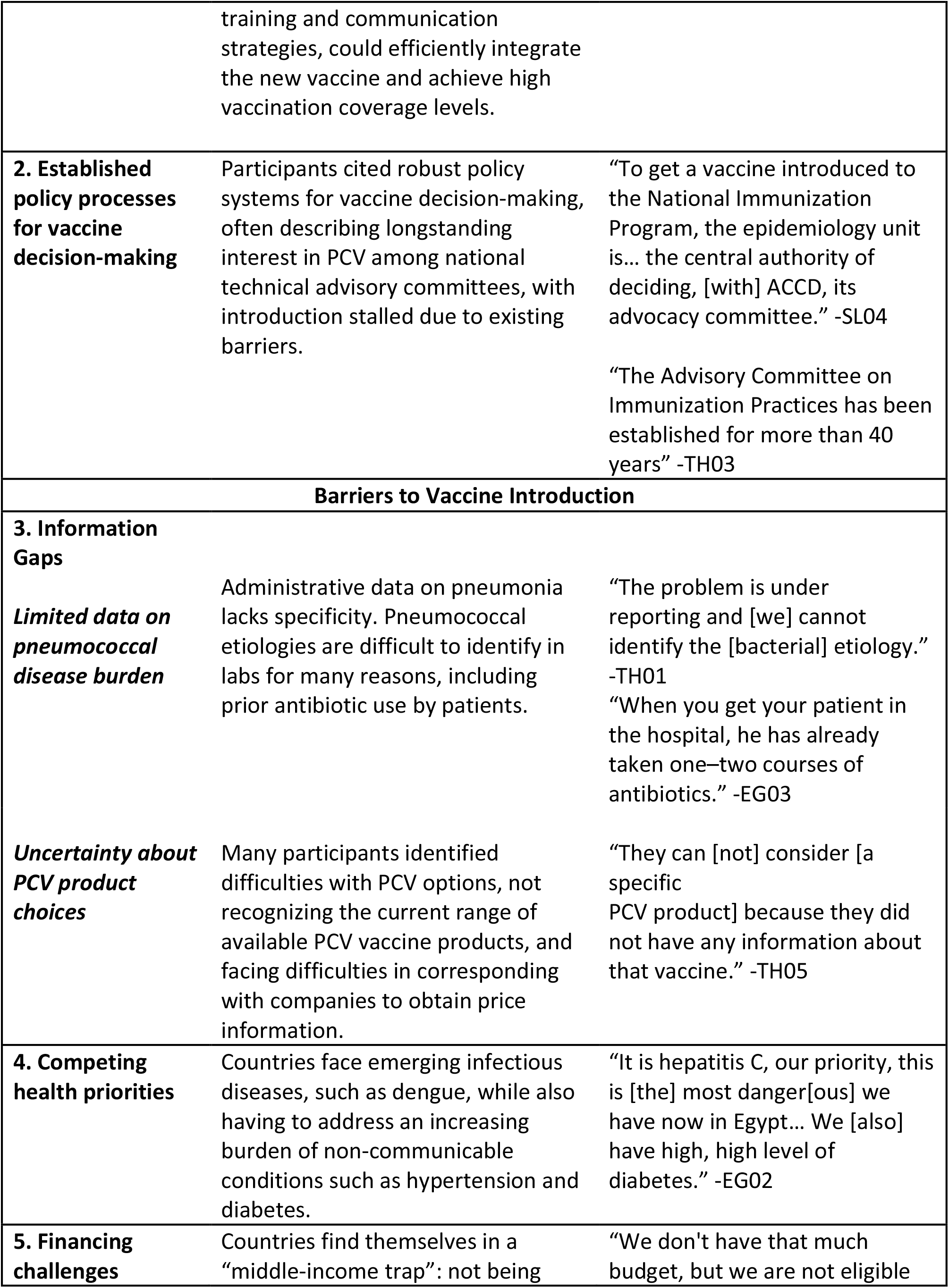
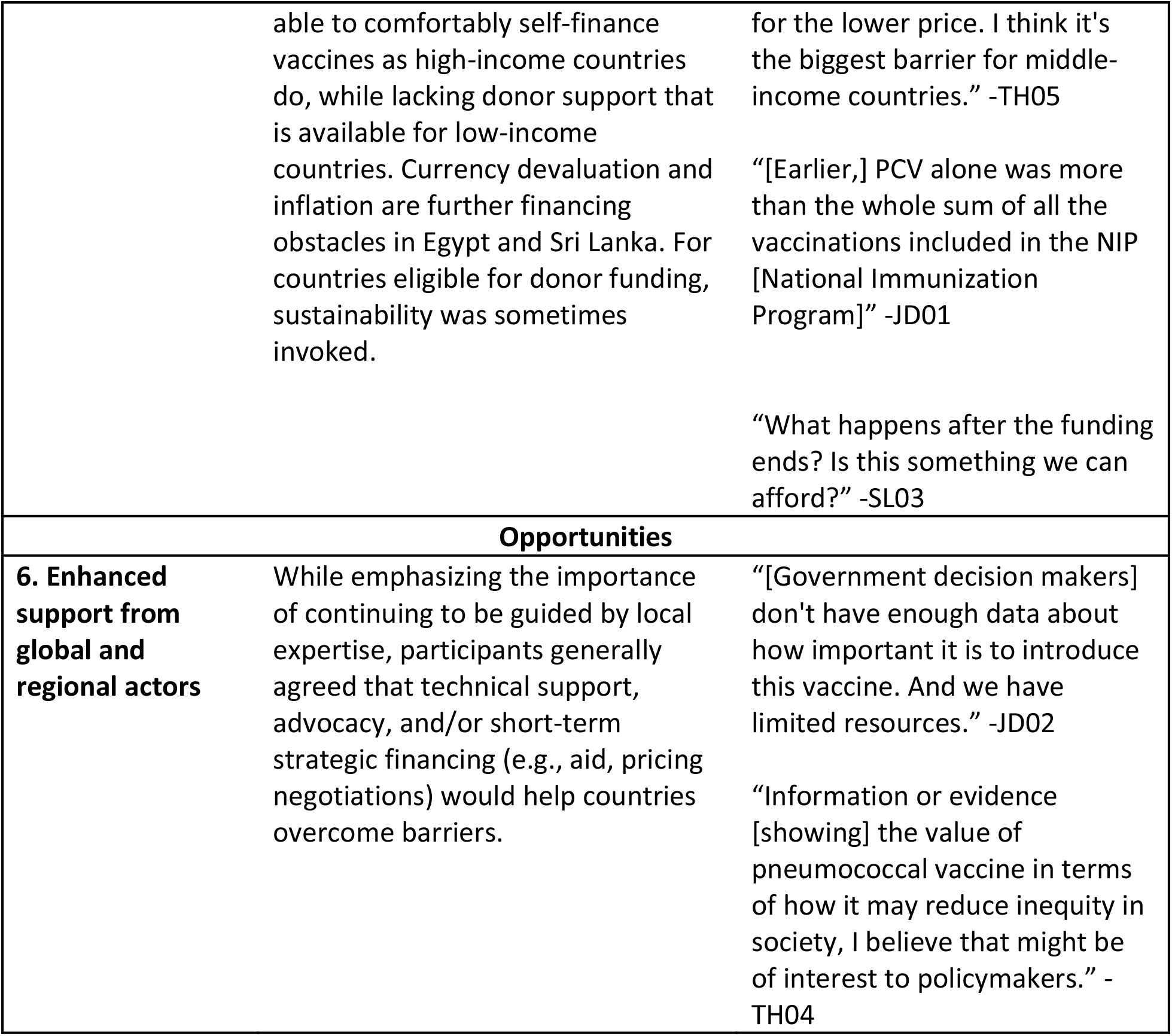
Thematic Issues Related to PCV Introduction in Four Middle-Income Countries (Egypt, Jordan, Sri Lanka, and Thailand)

| Themes | Description | Quotes |
| --- | --- | --- |
| **Factors Enabling Vaccine Introduction** |  |  |
| **1. Strong primary healthcare systems** | Should these countries choose to introduce PCV, their strong health systems, complemented by robust | "The implementation of PCV would be smooth if the Ministry made the decision." -EG04 |

<sup>1</sup> 16 contacted in Egypt, four in Jordan; 16 Thailand; 17 in Sri Lanka

|  |  |  |
| --- | --- | --- |
|  | training and communication strategies, could efficiently integrate the new vaccine and achieve high vaccination coverage levels. |  |
| **2. Established policy processes for vaccine decision-making** | Participants cited robust policy systems for vaccine decision-making, often describing longstanding interest in PCV among national technical advisory committees, with introduction stalled due to existing barriers. | “To get a vaccine introduced to the National Immunization Program, the epidemiology unit is... the central authority of deciding, [with] ACCD, its advocacy committee.” -SL04<br>“The Advisory Committee on Immunization Practices has been established for more than 40 years” -TH03 |
| **Barriers to Vaccine Introduction** |  |  |
| **3. Information Gaps** |  |  |
| *Limited data on pneumococcal disease burden* | Administrative data on pneumonia lacks specificity. Pneumococcal etiologies are difficult to identify in labs for many reasons, including prior antibiotic use by patients. | “The problem is under reporting and [we] cannot identify the [bacterial] etiology.” -TH01<br>“When you get your patient in the hospital, he has already taken one–two courses of antibiotics.” -EG03 |
| *Uncertainty about PCV product choices* | Many participants identified difficulties with PCV options, not recognizing the current range of available PCV vaccine products, and facing difficulties in corresponding with companies to obtain price information. | “They can [not] consider [a specific PCV product] because they did not have any information about that vaccine.” -TH05 |
| **4. Competing health priorities** | Countries face emerging infectious diseases, such as dengue, while also having to address an increasing burden of non-communicable conditions such as hypertension and diabetes. | “It is hepatitis C, our priority, this is [the] most danger[ous] we have now in Egypt... We [also] have high, high level of diabetes.” -EG02 |
| **5. Financing challenges** | Countries find themselves in a “middle-income trap”: not being | “We don't have that much budget, but we are not eligible |
|  | able to comfortably self-finance vaccines as high-income countries do, while lacking donor support that is available for low-income countries. Currency devaluation and inflation are further financing obstacles in Egypt and Sri Lanka. For countries eligible for donor funding, sustainability was sometimes invoked. | for the lower price. I think it's the biggest barrier for middle-income countries." -TH05<br>"[Earlier,] PCV alone was more than the whole sum of all the vaccinations included in the NIP [National Immunization Program]" -JD01<br>"What happens after the funding ends? Is this something we can afford?" -SL03 |
| **Opportunities** |  |  |
| **6. Enhanced support from global and regional actors** | While emphasizing the importance of continuing to be guided by local expertise, participants generally agreed that technical support, advocacy, and/or short-term strategic financing (e.g., aid, pricing negotiations) would help countries overcome barriers. | "[Government decision makers] don't have enough data about how important it is to introduce this vaccine. And we have limited resources." -JD02<br>"Information or evidence [showing] the value of pneumococcal vaccine in terms of how it may reduce inequity in society, I believe that might be of interest to policymakers." -TH04 |

### Factors enabling PCV introduction

#### Strong primary health care systems

Participants emphasized health systems strengths and universally agreed that, given a policy decision to introduce PCV in their countries, the implementation would be smooth and successful. Newly introduced vaccines would soon reach the same high coverage levels as other vaccines already in the routine system. Participants from Sri Lanka discussed the country’s experience maintaining high coverage even during periods of internal conflict and, after a brief dip in 2020, throughout the COVID-19 pandemic. One pediatric leader commented on the country’s program success:

> *“The vaccine coverage for primary immunization schedule [has been] near a hundred percent for at least five years. The maintenance of cold chain is strictly adhered to. And there’s a good support system for vaccine related adverse effects by pediatricians who are always supportive of the vaccination programs*.*”* –SL04

Participants shared that such high coverage levels could be attributed to high levels of trust and support for vaccination, as well as strong communication strategies to ensure high demand for new vaccines. In Egypt and Thailand, participants discussed a standard approach to developing health communications campaign packages for use with parents to proactively address potential concerns for a new vaccine like PCV. In Jordan, participants also described historically strong coverage rates. However, Jordanian participants also shared that there has been recent COVID-19-driven erosion in vaccine confidence. Despite this trend, participants felt that effective communication campaigns during implementation would address PCV-related hesitancy.

A caveat to the depiction of well-managed health systems was the current inequality in access to vaccines available only in the private market. Across the four countries, participants shared that the high cost of PCV was contributing to health inequities as PCV was only available to those who could afford to pay out-of-pocket.

> *“It is apparent that the high-income people and those who are well educated are keen to have their kids vaccinated*.*” – EG02*

#### Established policy processes for vaccine decision-making

Participants described historically well-organized policy-making structures that balance urgent health priorities, as well as economic and humanitarian crises, with equity-minded purchasing decisions. Participants generally concluded that new vaccines would be easily integrated into existing systems due to their robust decision-making capacity. However, in some countries, participants shared that processes were altered by a rapid push to introduce COVID-19 vaccines. Further, the economic crisis, arriving on the heels of the COVID-19 pandemic in Sri Lanka, has affected the prioritization of PCV.

> “*This advisory committee was again promoting pneumococcal vaccine [in 2020]. This collapsed after that, after the [economic] trouble after the COVID-19*.” -SL02.

Similarly, disruptions in NITAG functioning, specifically the turnover in its expert membership, have delayed PCV introduction in Jordan. Further, policy processes, specifically the pilot programs for introducing new vaccines, have delayed introductions of new vaccines in Thailand. These ‘pilot studies’, which are implemented ahead of potential national vaccine rollouts, have been reported to set back vaccine introduction by up to a decade in the case of rotavirus vaccine introduction; this is also delaying a potential PCV introduction in the country.

### Barriers to introduction

#### Information gaps

Participants cited information gaps as a leading barrier to PCV introduction. These gaps were clustered into the following two categories: 1) data on childhood pneumonia and pneumococcal disease; and 2) uncertainty about PCV product choices.

#### Limited data on pneumococcal disease burden

Across the four countries, participants described child pneumonia as being low in political prominence due to a limited understanding of the etiology of severe pneumonia cases presenting at hospitals, and strong access to facility-based treatment, preventing pneumonia mortality. Consequently, participants shared that the burden of pneumococcal disease was not well understood, and in most cases, they felt it was underestimated.

Although there were limited sources of data about invasive bacterial disease, participants were often aware of high community carriage rates, with several participants citing specific estimates of local disease burden generated through carriage studies. Participants further emphasized limited studies showing that existing vaccine products will offer strong coverage against circulating serotypes. However, the four countries do not routinely use serotyping to monitor circulating strains.

Participants’ understanding of the disease burden is further obstructed by the absence or inconsistency of administrative data on hospitalized pneumonia cases. This lack of data sharply contrasts with the more robust reporting systems in place for other infectious diseases.

> “We don’t have a database for pneumococcal. This is one of the major problems. In Jordan we have 42 diseases [that we have surveillance for], but pneumococcal [is] not one of them.”-JD02

The widespread use of antibiotics at the community or primary level was noted as a key reason the pneumococcus was infrequently detected in laboratory settings.

> *“We’ve definitely found positive tests for… pneumococcal, but very low incidence. The problem is the antibiotic use before admission*.*” -* Thailand, TH01
>
> *“We are a tertiary care hospital, [so] our lab does not yield a large number of pneumococcal isolates [where] patients have been visiting the primary and secondary care,” -*EG03

In Thailand and Sri Lanka, participants emphasized their low child mortality rates. Further, participants from Sri Lanka suggested that investments in PCV introduction would have a low return-on-investment compared to government programs to address neonatal causes of mortality.

“*Having such a low, single-digit [under 5] mortality rate, most of it [caused by] neonatal problems and birth defects… Even if we start a campaign of providing pneumococcal vaccination, we aren’t quite sure [this compared well with] money spent with neonatal services and community-based services*.*”* -SL05

However, some participants felt that a lack of surveillance data should not stop the government from introducing PCV. Across all four settings, participants agreed that a perceived lack of data has led to a delay or lack of prioritization of PCV introduction, but this lack of data was not indicative of a lack of disease burden.

> *“We don’t have a lot of children die from pneumonia, but we do have a lot of children come down with pneumonia. That is avoidable*.*”* -TH02

#### Uncertainty about PCV product choices

Participants were unfamiliar with the current range of PCV products and discussed resultant challenges in vaccine procurement.

Several participants were unfamiliar with lower priced PCV products. Policy discussions about specific PCV products, which country leaders felt hadn’t been sufficiently tested in diverse contexts, were reported by participants as impacting decision-making processes. In Thailand, several participants described vaccine procurement negotiations as also contributing to slowing vaccine decision-making processes. They commented that the tiered pricing available to MICs like Thailand was higher than the PAHO pooled price for PCV. Vaccine procurement in Jordan was reported to be complicated due to population dynamics. As Jordan is currently navigating an influx of refugees, accessing external support for vaccine programs was challenging. A Jordanian participant further said the cold chain may need to be expanded and that storage-related limitations may drive product choices:

> *“All these aspects [determine] the decision on which type of vaccine: how many serotypes included, even the size of the vaccine and [how many can fit in the] cold room*.*” JD03*

#### Competing health priorities

The second barrier to introduction was competing country health priorities for funding, including new emerging infectious diseases and non-communicable diseases (NCDs).

Participants reported that there has been advocacy and “*political pressure*” to prioritize and introduce other vaccines, including HPV and, depending on approvals and availability, dengue vaccines. In both Jordan and Sri Lanka, participants also described COVID-19 as a new “*seasonal disease*” that is still killing people.

> *“When you have so many competing priorities, particularly with COVID-19, there’s no money left*.*” -*TH02

Meanwhile, all four countries have seen an increase in the burden of NCDs, with countries reporting the need to intervene to address nutritional health, obesity, and a rising burden of diabetes.

Participants in both Jordan and Egypt described humanitarian crises and an influx of refugees, with large child populations in need of health services, as diverting political attention from new vaccine introductions.

> *“It [PCV] is a priority, but there [were] emergencies, and they couldn’t take action. The Sudan response is making things [get] postponed*.*” EG04*

Finally, there are other types of emerging priorities, such as injuries, as seen in Thailand, where participants shared that they are facing a growing burden of road accidents.

#### Financing

The high price of PCV was the third and most salient barrier discussed, with participants from all four countries citing funding concerns as delaying PCV introduction.

In Jordan, the NITAG first recommended introducing PCV in 2011, along with three other vaccines. While Jordan has introduced Hepatitis A and Rotavirus, they have not yet introduced PCV. A participant described extensive negotiations and the need to take a long-term perspective when advocating for vaccine introduction.

> *“After five or 10 years [PCV] will save money by decreasing the prevalence of pneumococcal diseases in Jordan, decreasing admissions due to pneumococcal diseases. But the mindset of the financial person is, if I go forward to introduce the PCV, I have to pay one third of the current [EPI] budget to introduce one vaccine*.*”-JD03*

In Egypt, participants similarly emphasized financial barriers as the most important obstacle to PCV introduction, stating that the government intends to introduce the vaccine but has been stymied by its economic crisis.

Although Thailand’s Technical Advisory Group previously recommended the introduction of PCV, the cost of PCV was described as the biggest barrier to vaccine introduction.

> *“That’s the biggest problem of the vaccine in Thailand. [Our vaccine] cost is similar to [a] high income country, but we are just an upper middle-income country. We [are] resource constrained, but [we are not] eligible for Gavi*.*” TH-05*

Participants also described a financial paradox or “trap” unique to MICs. These countries are not able to comfortably self-finance vaccines as in high-income countries, but they are also not able to access donor support to introduce them as in LICs.

> *“[As a] middle income country we’re trapped. We have difficulties to access vaccines compared to our colleagues in neighboring countries. Our program has acknowledged about the delay compared to other countries, even compared to Gavi countries*.*” – TH03*

Participants also described country-specific barriers associated with PCV pricing. Participants in Egypt and Sri Lanka described precarious economies with currency devaluations affecting vaccine decisions. In Egypt, respondents shared that the government was seeking to manufacture PCV in-country, in part to bypass the currency exchange problem. However, a major barrier to this, as cited by participant EG02, was that “*every process will need some [supplies] to be imported from outside*.”

Similarly, in Sri Lanka, a pediatric leader, SL05, said “*because of the [currency] difficulties, the government had to limit imports very drastically*.*”*

### Opportunities for introduction

#### External support

A key opportunity described by participants was the effect of potential support from entities outside the government, for instance, from regional or global actors, in the form of technical assistance, advocacy materials, or subsidies.

Participants described distinct roles for external support. In Sri Lanka, some participants felt that with Gavi support through the MICs Approach they could introduce and later self-sustain PCV. However, other participants had questions about long-term pricing following the infusion of donor support.

Participants also described receiving external technical advice to understand where they stand in relation to other countries’ vaccine introduction status was a motivating factor. A participant from Jordan referred to an internal meeting where a regional map of vaccine introduction, provided by an external group, was shown.

> *“Honestly, the map comparing Jordan to neighboring countries helped a lot. Because when you look at the map [of] the region and see the countries that introduced PCV, [you] see that [Jordan] is still lagging behind a lot. Jordan is supposed to be one of the health pioneers in the region*.*” -JD04*

Similarly, participants in Thailand described the country’s sense of its Expanded Program on Immunization (EPI) schedule as “*not very advanced*” compared to neighboring countries. They saw a further role for global actors; frustrated with high prices from global suppliers that some participants found “*unethical*,” one participant suggested a third party, for instance, a donor agency, could negotiate for Thailand to receive a certain price to expedite vaccine introduction.

Participants also cited a need for collective action, with most saying that support from influential scientific leaders and external groups is essential to new introductions.

> *“When it comes from WHO’s side, it comes from UNICEF, it comes from [academic institutions], or wherever. That’s [when] you know that there’s something bigger going on. It’s unnatural that all those entities [would] agree on certain things unless there’s a real benefit*.*” -JD04*

## Discussion

Our interviews with health leaders in Egypt, Jordan, Sri Lanka, and Thailand revealed key insights on contextual factors to support PCV introduction in MICs, including leveraging robust primary healthcare systems and policy processes for vaccine decision-making. Notable challenges included limited information on pneumococcal disease burden and vaccine products, competing health priorities, and financing considerations. Importantly, participants identified opportunities to support new vaccine introductions through strategic financial support, technical assistance, and advocacy from global and regional actors. In fact, in a promising development, since Jordanian leaders were engaged for this study, Jordan announced plans to introduce PCV in mid-2024 with support from Gavi’s MICs Approach[19], [20].

While recent global policy briefs and major vaccine donors have highlighted the lagging pace of vaccine introductions and the large burden of vaccine-preventable deaths in MICs, this paper fills an evidence gap by qualitatively exploring the contextual reasons why PCV has not yet been implemented into the routine immunization system of four MICs [8], [21].

Our results complement a MICs strategy briefing shared with the Strategic Advisory Group of Experts on Immunization (SAGE) in 2015, which highlighted four bottlenecks in MICs: decision-making, financial sustainability, poor demand and underperforming services, and affordable access to supplies[22]. While these factors are likely still present in a variety of MIC contexts, our respondents emphasized challenges around just two of the four bottlenecks. Our participants shared that their respective countries have well-established policy processes, as well as high population demand and quality services.

Respondents described strong immunization programs with high levels of population trust and robust coverage. While MICs that are ineligible for traditional Gavi support suffered the most COVID-19 pandemic backsliding of any group of countries, coverage for key routine vaccines in Egypt, Sri Lanka, and Thailand remained consistently high from 2019–2022[23], [24]. Such remarkable resiliency reinforces participants’ statements that implementing a new vaccine, such as PCV, even with some concurrent financial and humanitarian challenges, can be swift and successful.

The dearth of data and perceived low disease burden are both linked to the low laboratory yield of pneumococcus; this has consistently hindered PCV introduction. While increased access to antibiotics has, fortunately, made pneumonia less fatal for children, it has also contributed to reduced detection of pneumococcus in the laboratory[25]. Despite WHO’s recommendations for active and passive surveillance in both high- and low-income countries, MICs have limited surveillance networks, which further reduce the visibility of the countries’ pneumococcal disease burden [26]. Because of this limited surveillance and subsequent lack of data, childhood pneumonia and pneumococcal disease may not be seen as urgent threats to policymakers in MICs.

Additionally, many countries are going through an epidemiological transition as they continue to see low child mortality rates and an increasing burden of NCDs, impacting the decision-making process for introducing new childhood vaccines. The growing population of middle-aged and older individuals highlights the urgent need for large-scale interventions to address NCDs, such as hypertension and diabetes. Simultaneously, leaders in MICs, particularly those in regions experiencing humanitarian crises, face the significant challenge of providing care to over half of the world’s refugees. This places additional demands on already strained health services, highlighting the need for comprehensive support and resources [27]. The combination of these factors, as well as the epidemiological transition these countries are undergoing, further complicates the decision-making process for childhood vaccines [28].

Information gaps also extend to a lack of awareness of newer PCV products, those that may be available at a lower cost but with comparable quality and efficacy[29]. Participants correctly identified that when vaccines are manufactured by a few companies in high-income settings, sustainable access in other settings is fraught. Bottlenecks to sustainably sourcing COVID-19 vaccines have prompted new initiatives to broaden vaccine manufacturing, such as the US $1 billion African Vaccine Manufacturing Accelerator; this initiative aims to support sustainable manufacturing for vaccines, including producing a minimum 13-valent PCV in Africa[30], [31]. Simultaneously in Asia, Thailand has recently approved its first domestically produced COVID-19 vaccine, and advocates have highlighted the strong potential for regional vaccine manufacturing in neighboring countries[32], [33]. These approaches can be leveraged to find innovative approaches to support PCV introduction in MICs.

Financing challenges, a significant barrier reported in our study, were also documented as a significant bottleneck in 2015[22]. Despite the evolving landscape of technical and financial support for MICs, participants in our study described feeling “trapped,” “delayed,” and “lagging” due to the ‘paradox’ we identified in our results, in which MICs were not able to comfortably self-finance vaccines nor able to access donor support to introduce vaccines. Vaccine financing has long been known as a critical constraint to PCV introduction. Gavi’s MICs Approach promises to promote further introductions of new and underutilized vaccines in eligible countries. Further, in a major development, UNICEF recently published revised PCV pricing for MICs, starting at $2.90 per dose[34]. Countries can also explore diverse innovative options for financing vaccines, including earmarking, domestic trust funds, strategic purchasing, and procurement options like pooled procurement, including through UNICEF procurement and financing mechanisms[35]. Building on the success of UNICEF’s Vaccine Independence Initiative, the MICs Financing Facility (MFF) will allow countries to benefit from UNICEF’s procurement, scale, access, market expertise, and affordable pricing[36].

Most participants also flagged a need for more diverse sources of support—technical assistance, advocacy, or short-term financial support. Targeted assistance will need to be responsive to expressed country interests and needs and provided expeditiously to address complex health systems challenges.

There were some limitations to the study. We recruited a smaller number of participants from government agencies in two of the four countries. Deeper participation from the countries’ governments and health ministries could have provided a more rounded understanding of the issues as well as potential solutions. However, all participants were deeply familiar with the health systems, policymaking processes, and introduction constraints. In-depth interview methodologies are prone to recall bias in that participants without an existing interest in PCV advocacy may have overlooked important policy procedures relevant to vaccine decision-making. The perspectives described were co-constructed in conversations between U.S.-based researchers and ‘elite’ health systems leaders and political influencers from MICs. Due to the nature of qualitative research conducted across borders, participants may have been reluctant to fully reveal some aspects of the immunization decision-making context. Finally, the participants’ perspectives reflect a correct understanding of global vaccine pricing for MICs prior to changes facilitated through Gavi’s MICs Approach, developments briefly described in the Discussion section that may continue to evolve with subsequent policy revisions.

## Conclusion

While Egypt, Jordan, Sri Lanka, and Thailand have developed strong, equity-focused health systems, implementing new vaccines such as PCV in middle-income settings has been challenging for a range of reasons, most notably sustainable vaccine financing. MIC have faced particularly unique challenges in the vaccine procurement process, as they were not able to comfortably self-finance vaccines for their birth cohort, but most have historically not been able to access donor support for new introductions. It is essential to leverage the current strengths of MICs, including existing health systems and robust policy procedures, to ease the introduction process. Global and regional partners must go further to support MICs in procuring vaccines thoughtfully and constructively, particularly for countries who are not eligible for donor support, in order to overcome inequalities in vaccine access. Accelerating this process in MICs where the majority of the world’s low-income and many high-need populations reside offers a means for reaching all children with life-saving immunization services and working towards strengthening global vaccine equality.

## Data Availability

Excerpts of the transcripts relevant to the study are available upon reasonable request to the authors.

## Acknowledgements

We are grateful to the participants who, among many competing national priorities, graciously shared their time and expertise with our interviewers. We also thank Sophie La Vincente, Meredith Shirey, Anna Osborne, and Caitlin Longden for their support in interpreting revised policy support from Gavi, the Vaccine Alliance for middle-income countries.

## AUTHOR CONTRIBUTIONS

BD and AS conceptualized the study. RW, BD, JH, and SN facilitated data collection and research activities. RW, BD, and AS drafted the manuscript, with JH, SN, AA, KC, LF, and EB interpreting and contextualizing the data and presentation of results. All authors reviewed, read, and approved the final manuscript and agree to be accountable for all aspects of the work. All authors declare that they have no competing interests.

## Footnotes

1 16 contacted in Egypt, four in Jordan; 16 Thailand; 17 in Sri Lanka

